# Comparing COVID-19 vaccine allocation strategies in India: a mathematical modelling study

**DOI:** 10.1101/2020.11.22.20236091

**Authors:** Brody H Foy, Brian Wahl, Kayur Mehta, Anita Shet, Gautam I Menon, Carl Britto

## Abstract

**Background:** The development and widespread use of an effective SARS-CoV-2 vaccine could help prevent substantial morbidity and mortality associated with COVID-19 infection and mitigate many of the secondary effects associated with non-pharmaceutical interventions. The limited availability of an effective and licensed vaccine will task policymakers around the world, including in India, with decisions regarding optimal vaccine allocation strategies. Using mathematical modelling we aimed to assess the impact of different age-specific COVID-19 vaccine allocation strategies within India on SARS CoV-2-related mortality and infection.

**Methods:** We used an age-structured, expanded SEIR model with social contact matrices to assess different age-specific vaccine allocation strategies in India. We used state-specific age structures and disease transmission coefficients estimated from confirmed Indian incident cases of COVID-19 between 28 January and 31 August 2020. Simulations were used to investigate the relative reduction in mortality and morbidity of vaccinate allocation strategies based on prioritizing different age groups, and the interactions of these strategies with several concurrent non-pharmacologic interventions (i.e., social distancing, mandated masks, lockdowns). Given the uncertainty associated with current COVID-19 vaccine development, we also varied several vaccine characteristics (i.e., efficacy, type of immunity conferred, and rollout speed) in the modelling simulations.

**Results:** In nearly all scenarios, prioritizing COVID-19 vaccine allocation for older populations (i.e., >60yrs old) led to the greatest relative reduction in deaths, regardless of vaccine efficacy, control measures, rollout speed, or immunity dynamics. However, preferential vaccination of this target group often produced higher total symptomatic infection counts and more pronounced estimates of peak incidence than strategies which targeted younger adults (i.e., 20-40yrs or 40-60yrs) or the general population irrespective of age. Vaccine efficacy, immunity type, target coverage and rollout speed all significantly influenced overall strategy effectiveness, with the time taken to reach target coverage significantly affecting the relative mortality benefit comparative to no vaccination.

**Conclusions:** Our findings support global recommendations to prioritize COVID-19 vaccine allocation for older age groups. Including younger adults in the prioritisation group can reduce overall infection rates, although this benefit was countered by the larger mortality rates in older populations. Ultimately an optimal vaccine allocation strategy will depend on vaccine characteristics, strength of concurrent non-pharmaceutical interventions, and region-specific goals such as reducing mortality, morbidity, or peak incidence.

## INTRODUCTION

After first emerging in Wuhan, China in late 2019,(Li et al. 2020) severe acute respiratory syndrome coronavirus 2 (SARS-CoV-2), the virus that causes coronavirus disease 2019 (COVID-19), has spread rapidly throughout the world causing local epidemics in virtually all countries (Dong et al. 2020). The virus is spread primarily through respiratory droplets and close contact with infected individuals (Chan et al. 2020; Li et al. 2020). While early, large-scale COVID-19 epidemics occurred primarily in high-income countries in Europe and North America, a significant number of cases and deaths attributable to the virus have also now occurred in low- and middle-income countries (Dong et al. 2020). Approximately 1.4 billion people are at risk of SARS-CoV-2 infection in India with many having risk factors for severe outcomes (Nandi et al. 2020).

The first confirmed case of COVID-19 in India was identified in Kerala on 30 January, 2020 in an Indian national who returned from China (Perappadan 2020). During the following month-and-a-half, several travel-associated cases were confirmed throughout the country (Rawat 2020). To slow the spread of the virus and reduce peak incidence, the central government initiated one of the largest lockdowns in the world on 25 March 2020 comprising several non-pharmaceutical interventions (NPIs), including physical distancing and restrictions on non-essential travel (Pulla 2020). Several analyses have indicated that lockdown measures reduced the effective reproduction number (*R*_*t*_) in the country (Gupta et al. 2020; Kumar et al. 2020; Sardar et al. 2020). However, the lockdown precipitated several secondary effects, including reduced livelihoods, difficulties with accessing routine health services, and mass migrations (*The Lancet* 2020). Lockdown measures were relaxed beginning on 1 June 2020 and as of 12 November 2020, India has reported approximately 8.6 million cases and 130,000 deaths (Dong et al. 2020).

In the absence of a highly effective therapeutic agent for COVID-19, the development of vaccines that provide protection from SARS-CoV-2 infection is a global imperative. An unprecedented effort is currently underway to rapidly develop effective COVID-19 vaccines, with several stakeholders working together to compress a process that normally takes years or decades into months (Graham 2020). Of the numerous vaccine candidates currently under development, 11 are in Phase III clinical trials as of November 2020 (Milken Institute 2020). Five COVID-19 vaccines have been approved for limited use; however, Phase III clinical trial data are not yet available for these vaccines and they have not been prequalified by the World Health Organization (WHO) (Milken Institute 2020). In India, clinical development is currently underway for multiple candidate vaccines.

Concurrent with the clinical testing of these vaccine candidates, new mechanisms are being established to expedite manufacturing and deployment and support the fair distribution of COVID-19 vaccines once they are licensed (World Health Organization 2020a). COVAX is a collaborative venture that was launched in June 2020 as the vaccine pillar of the Access to COVID-19 Tools (ACT) Accelerator (World Health Organization 2020a). An important component of this venture is the COVAX Facility, a global risk-sharing mechanism for the pooled procurement of COVID-19 vaccines. Through this mechanism, 92 low- and middle-income countries, including India, are eligible to be supported by the COVAX Advance Market Commitment (AMC), which will pay for the cost of COVID-19 vaccines once COVID-19 vaccines have been licensed and prequalified by the WHO.

Based on WHO guidelines and recommendations related to which priority groups should receive COVID-19 vaccines first (World Health Organization 2020b), countries participating in the COVAX Facility are encouraged to prioritize frontline health workers and social care workers. Toward that end, the mechanism aims to distribute COVID-19 vaccines equally to countries until they have achieved a sufficient supply to protect this vulnerable population, determined to represent approximately 3% of country populations. As the supply of COVID-19 vaccines increases, the distribution of COVID-19 vaccines through the COVAX Facility will continue such that 20% of country populations can be covered, specifically those at increased risk (i.e., older adults and those who have underlying health conditions).

In the context of limited supply and to support policies related to COVID-19 vaccine allocation in India, we developed a mathematical model to simulate different vaccine allocation strategies. There remain several unknowns associated with the current COVID-19 vaccine development. Therefore, we assessed these vaccine allocation strategies varying potential vaccine characteristics. We also evaluated the relative reduction in cases and deaths in the context of varying control measures (NPIs). The findings of this analysis could also be used by other low- and middle-income countries to inform their COVID-19 vaccine allocation strategies.

## METHODS

### Data Collection

Daily and state-specific incident SARS-CoV-2 infection case data were collected from multiple sources, including the Ministry of Health and Family Welfare (MOHFW), the Indian Council of Medical Research (ICMR), and a website for crowd-sourced information related to COVID-19 in India (www.covid19india.com). The data available from this website are collated from public sources and validated by a group of volunteers.

### Model of disease transmission

Disease transmission in Indian populations was modelled using an age-structured compartment model, stratified into ten-year age bands (0-10, 10-20, […], 60-70, ≥70 years). The model includes different compartments for each age band and infection state (i.e., *S, E, A, I, Q*, and *R*). We assume subjects start off susceptible to infection (*S*) and can become exposed (*E*) after contact with an infectious individual. After a latent period, exposed subjects either develop an asymptomatic (*A*) or symptomatic (*I*) infection, with an age-stratified probability. Subjects with symptomatic infections are either hospitalized or choose to self-isolate (*Q*) at a given rate. Once hospitalized or isolated, subjects either recover (*R*) or die (*D*), with an age-stratified mortality rate. Asymptomatic individuals are assumed to have no risk of mortality and simply recover at a given rate. Recovered subjects are assumed to become susceptible at a given rate, reflecting eventual loss of temporary immunity from the infection (Sariol and Perlman 2020). We assumed that COVID-19 vaccines are allocated gradually into a specific age-defined community at a constant rate.

We simulated two different mechanism through which COVID-19 vaccines could induce immunity (Peiris and Leung 2020). In one simulation, vaccinated individuals (*V*) are protected from infection and therefore unable to infect others if exposed to an infected individual (i.e., sterilizing immunity). In the other simulation, vaccinated individuals are not protected from asymptomatic infection if exposed and therefore can infect others if they become infected (i.e., non-sterilizing immunity). In the latter, if an individual develops an asymptomatic infection after receiving a vaccine that does not confer sterilizing immunity, they are assumed to have a temporary immunity from developing further asymptomatic infections, with immunity waning at the same rate as non-vaccinated subjects who recover from infection. Formulated as a system of differential equations, and using *S*_*i*_ to denote the susceptible population from age group *i*, for each age group our model comprises:

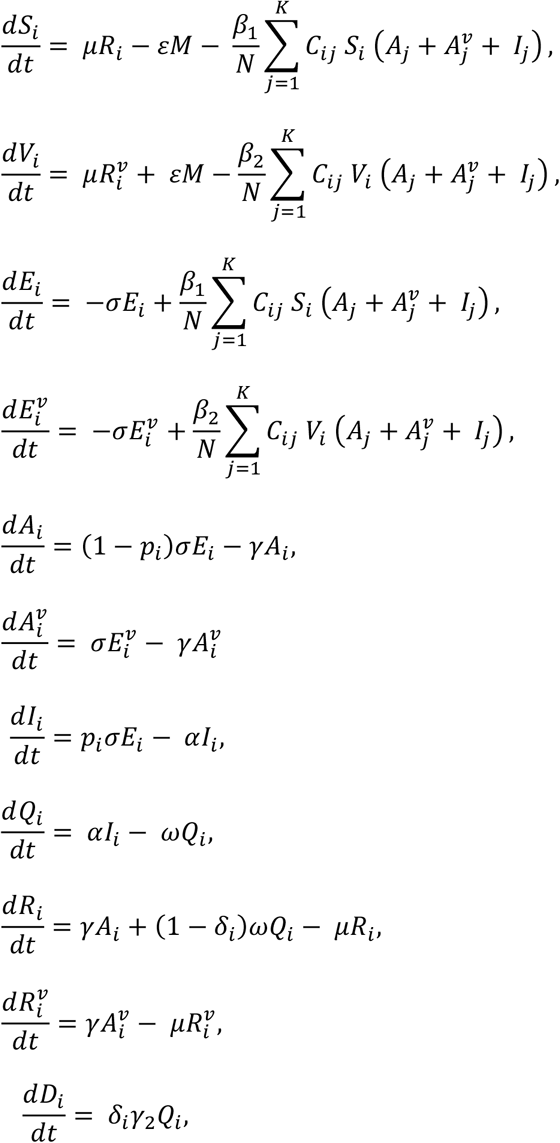

where 1/*μ* is the average length of immunity, *β*_1_ is the force of infection, *N* is the total population size, 1/σ is the average latent period, *p* is the proportion of infections which are symptomatic, 1/γ is the average asymptomatic recovery time, 1/ω is the average time from isolation to recovery for a symptomatic infection, 1/α is the average time until a symptomatically infectious subject self-quarantines or is hospitalized, and δ is the likelihood of death given symptomatic infection. *C*_ij_ is the relative frequency of contact between age group *i*and age group *j*. For the simulation where vaccines confer non-sterilizing immunity, *β*_2_ = *β*_1_ and *E*^*V*^, *A*^*V*^, and *R*^*V*^denote subjects who are exposed, asymptomatic, and recovered, respectively. For the simulation where COVID-19 vaccines provide sterilizing immunity, *β*_2_ = 0, meaning those who are vaccinated cannot become infectious. In both cases, vaccines are assumed to be rolled out gradually, with ε doses available each day, and an efficacy of ε. A flow diagram of transitions within the model is given in Figure 1.

**Figure 1:**
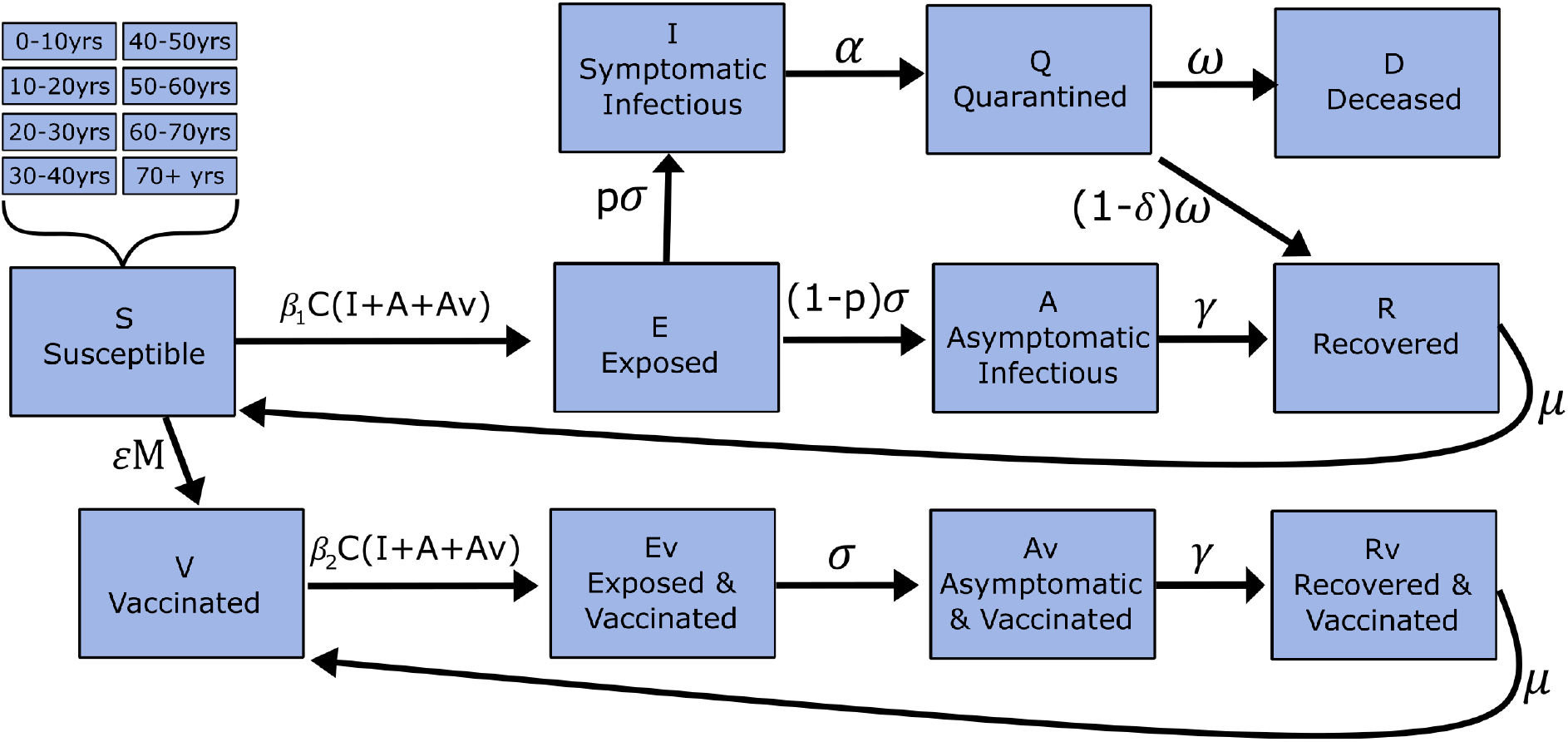
Schematic of model transmission dynamics. Subjects may move from susceptible to exposed to symptomatic or asymptomatic infectious. Asymptomatic infectious are assumed to always recover, while symptomatic infectious first quarantine, before recovering or dying and (if recovered) eventually losing immunity. Each major compartment comprises eight sub-compartments, comprising age groups (0-10, 10-20, […] 60-70, ≥70 years). Rates of symptomatic infection (*p*) and death (δ) vary by age group. Contact between susceptible and infectious populations is age-structured, proportional to the estimated contact pattern matrix (*C*). Under a progressive rollout scheme, ε individuals are vaccinated each week, at an efficacy of ε. If the vaccine confers non-sterilizing immunity, individuals can become exposed and develop asymptomatic infections, before recovering and eventually losing infection-driven immunity. For those vaccines that do not confer sterilizing immunity, *β*_2_ = 0; meaning vaccinated individuals no longer contribute to transmission dynamics.

Contact matrices (*C*) were estimated from social mixing patterns in the general Indian population (Prem et al. 2017). Estimates were broken down into four mixing categories, representing the four mixing patterns in different environments: (1) “at home”, (2) “at school”, (3) “at work”, and (4) “other”; with *C* representing the summation of the four mixing matrices. In normal scenarios (i.e., no control measures), each mixing pattern was equally weighted. Under strong control measures weights of 1.21, 0.56, 0.0, and 0.45 were used for “at home”, “at work”, “at school”, and “other” matrices respectively, based on estimates from Google’s mobility data during the lockdown period (March 25 – May 31, 2020) (Aktay et al. 2020). Moderate control measures were simulated using the average between no control and strong control measure weights.

All parameters except *β* were estimated based on prior studies, with a full list of parameters, estimated values, and their sources given in Table 1. *β* values were estimated based on fits of the model-simulated, hospitalized or self-isolated population numbers (*Q*) against confirmed active infection case numbers, between 1 June and 31 July (i.e., after the lockdown period). Given variability in social mixing patterns immediately after the national lockdown, *β* values were estimated assuming moderate control measures and no control measures during this period. Parameter fitting was performed using *MATLAB’s Statistical Toolbox* with an example data fit presented in the supplemental materials. All models were simulated in MATLAB and use forward Euler discretisation for the differential equations, with a timestep of one day.

**Table 1:** Model parameters and region-specific R_0_ estimates. R_0_ was estimated during the 2 months post-lockdown (June 01 – July 31, 2020), with either moderate or no non-pharmaceutical interventions (NPIs).

| **$R_0$ Estimates** |  |  |  |
| --- | --- | --- | --- |
| **Region** | **Population (millions)** | **Value**<br>**(no NPIs)** | **Value**<br>**(moderate NPIs)** |
| Uttar Pradesh | 204.2 | 2.41 | 3.22 |
| Maharashtra | 114 | 2.2 | 2.89 |
| Bihar | 104.1 | 2.36 | 3.15 |
| West Bengal | 90.3 | 2.33 | 3.10 |
| Madhya Pradesh | 72.6 | 1.81 | 2.39 |
| Tamil Nadu | 72.1 | 2.31 | 3.12 |
| Rajasthan | 68.5 | 2 | 2.59 |
| Karnataka | 64.1 | 3.74 | 4.95 |
| Gujarat | 60.4 | 1.89 | 2.49 |
| Andhra Pradesh | 49.5 | 2.93 | 3.92 |

| **Other model parameters** |  |  |
| --- | --- | --- |
| **Parameter** | **Value** | **Reference** |
| $1/\sigma$ (latent period) | 5.1 days | (Lauer et al. 2020) |
| $1/\gamma$ (recovery period) | 21 days | (Bi et al. 2020) |
| $1/\alpha$ (pre-isolation infection period) | 4.6 days | (Bi et al. 2020) |
| $1/\omega$ (post-isolation recovery period) | 16.4 days | (Bi et al. 2020) |
| $1/\mu$ (immunity duration) | 1 year | Estimated (other values in supplemental material) |
| $p$ (proportion of symptomatic infections) | Age-specific | (Davies et al. 2020) |
| $\Delta$ (case fatality rate) | Age-specific | (Laxminarayan et al. 2020) |

### Vaccination strategies

Four age-based vaccination strategies were considered: (1) vaccines are distributed evenly across the entire population or were first distributed to those who were: (2) 20-40 years, (3) 40-60 years, or (4) ≥60 years. In strategies 2-4, following vaccination of the target age group to the assumed vaccine coverage, vaccine doses were allocated evenly to the remaining population. Simulations were performed using a range of vaccine efficacies, and assuming a fixed number of doses available each day. For each age group, total vaccinations were set to not exceed the target coverage of the population, reflecting that many people will not be vaccinated, due to personal choice or local availability limitations.

Within this framework, simulations were performed using efficacy and target coverage ranging from 0-100% and considering vaccines that provide sterilizing and non-sterilizing immunity. Dose availability was assumed constant over time, reflecting the market pressures of acquiring vaccine doses, and the logistic pressures of distributing these doses. Dose availability was expressed as the percentage of the population which could be vaccinated each month, with simulations using values from 2-15%, reflecting an approximate time of between six months to four years to vaccinate to 100% of the target population.

## RESULTS

### Parameter estimates

To capture region specific infection dynamics, *R*_0_ values were estimated for the 10 most populous states within India, assuming moderate control measures and no control measures. Estimates are given in Table 1, with mean *R*_0_ values of 2.4 assuming no control measures and 3.2 assuming moderate control measures during the lockdown period. We used the former estimate as the base case value in our simulations. Minimum and maximum values were 1.8 and 5.0 respectively, with results using these values presented in the supplemental materials. The values in Table 1 are consistent with *R*_0_ values reported for other countries (Gatto et al. 2020; Sanche et al. 2020; Wu et al. 2020; Zhao et al. 2020). Within our model, the implementation of moderate and severe control measures led to a 23% and 44% relative reduction in *R*_0_, respectively.

### Vaccine strategy simulations

Four vaccine strategies were simulated under variations in dosage availability, target group coverage, vaccine efficacy, effect on transmission (i.e., sterilizing or non-sterilizing immunity), and the implementation of other control measures (i.e., no lockdown, moderate lockdown, or strong lockdown). Example epidemic curves for COVID-19 vaccines that confer sterilizing and non-sterilizing immunity are given in Figure 2. Regardless of vaccination strategy and immunization coverage in the target population, the initial infection wave occurs at a similar time, though with varying severity based on strategy. However, COVID-19 vaccines that confer sterilizing immunity appear to minimize the extent of future infection waves. In both cases strategy 4 (i.e., prioritizing individual ≥60 years) leads to the greatest reduction in deaths; however, all vaccination strategies produce significant benefits comparative to no vaccination.

**Figure 2:**
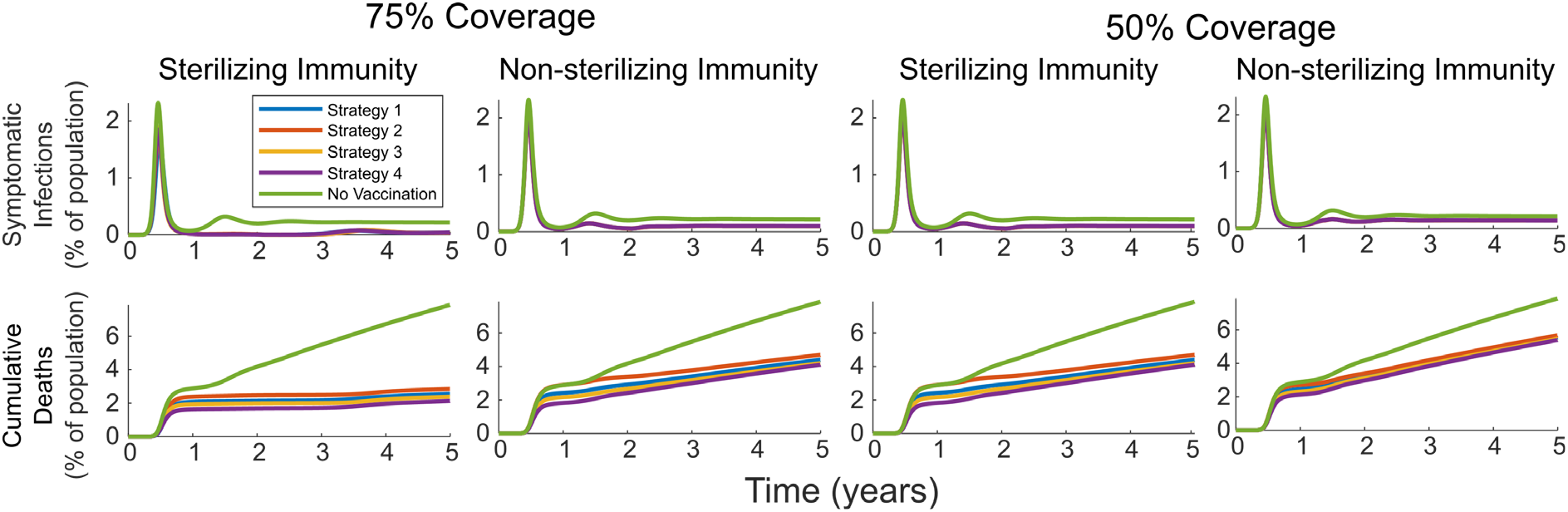
Simulated infection curves and cumulative deaths with four vaccination strategies. Each simulation assumed that 3% of the population was vaccinated each month, with a vaccine efficacy of 75%, no control measures, and an *R*_0_ of 2.4.

Within Figure 3, we present the estimated reduction in deaths and symptomatic infections over a five-year period using each of the four vaccination strategies, under varying efficacy, control measures, and rollout speeds. All results are presented relative to the outcomes with no vaccination, using the same *R*_0_ value, and with no control measures. Simulations were performed using an *R*_0_ of 2.4 (i.e., the mean *R*_0_ value in 10 states) and assume a target COVID-19 vaccine coverage of 75%. Results in Figure 3 illustrate that prioritizing vaccine allocation among older adults consistently results in the greatest reduction in deaths, regardless of vaccine efficacy, control measures, rollout speed, or immunity type. Conversely, all four strategies result in extremely similar reductions in symptomatic infection rates, with the optimal strategy being dependent on the specific implementation and vaccine.

**Figure 3:**
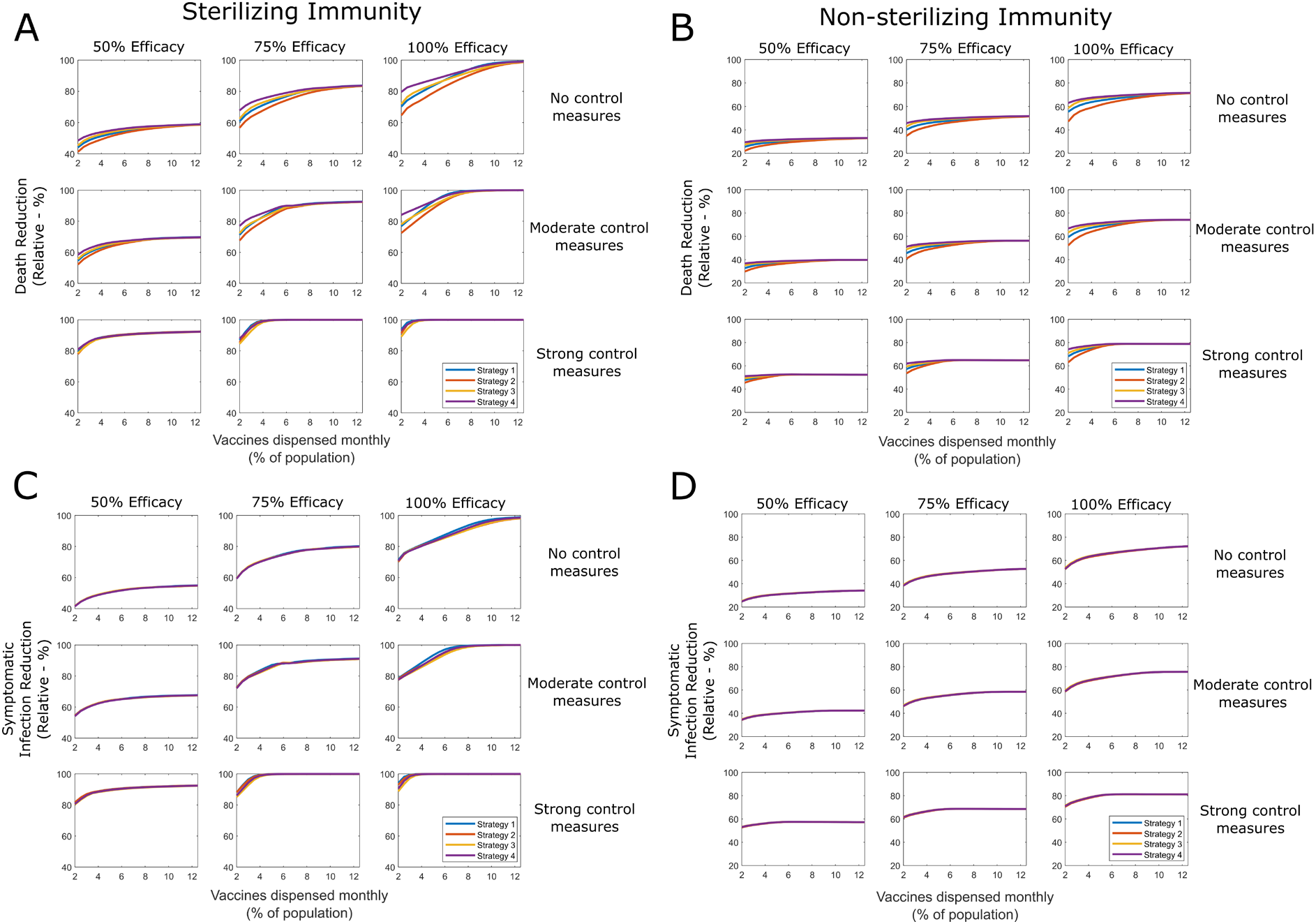
Comparison of benefit for four different vaccination strategies, against no vaccination. The relative reduction in deaths (A, B) and symptomatic infections (C, D) over a 5-year period are presented for four vaccination strategies, under varying speeds of vaccine dispensation. Results are stratified using three different vaccine efficacies, three types of control measure policy, and assuming a vaccine grants either sterilizing or non-sterilizing immunity. All simulations assumed vaccination did not exceed a population coverage of 75% and used an *R*_0_ of 2.4. Baseline deaths were calculated assuming no control measures and the same R0 value.

The relative benefit of prioritizing vaccine allocation among older adults compared to other strategies is highest under slower rollout speeds, while overall benefit is greatest the faster the rollout speed. Overall reduction in deaths is strongly limited by vaccine efficacy, and is strongly influenced by control measures, with more severe measures leading to greater reductions. Similar patterns were seen with different *R*_0_ values and target coverages (supplemental materials). Similar patterns were also seen when assuming imperfect self-isolation of symptomatically infectious individuals (supplemental materials).

While *R*_0_ values, vaccine efficacy, and other vaccine characteristics (i.e., sterilizing versus non-sterilizing immunity) all influence strategy effectiveness, in application these factors are immutable from the perspective of policy makers. Rather, international and national efforts, including investments and policies, can primarily influence three factors: (1) dosage availability/rollout speed, (2) target vaccine coverage; and (3) the continuation or relaxation of control measures. Within this context, in Figure 4 we present the relative reduction in deaths for vaccine allocation prioritizing older adults as each of those factors is modified. Equivalent results for strategies 1-3 are presented in the supplemental materials. Under low coverage, the speed at which the vaccine is rolled out has little effect on the overall reduction in deaths.

**Figure 4:**
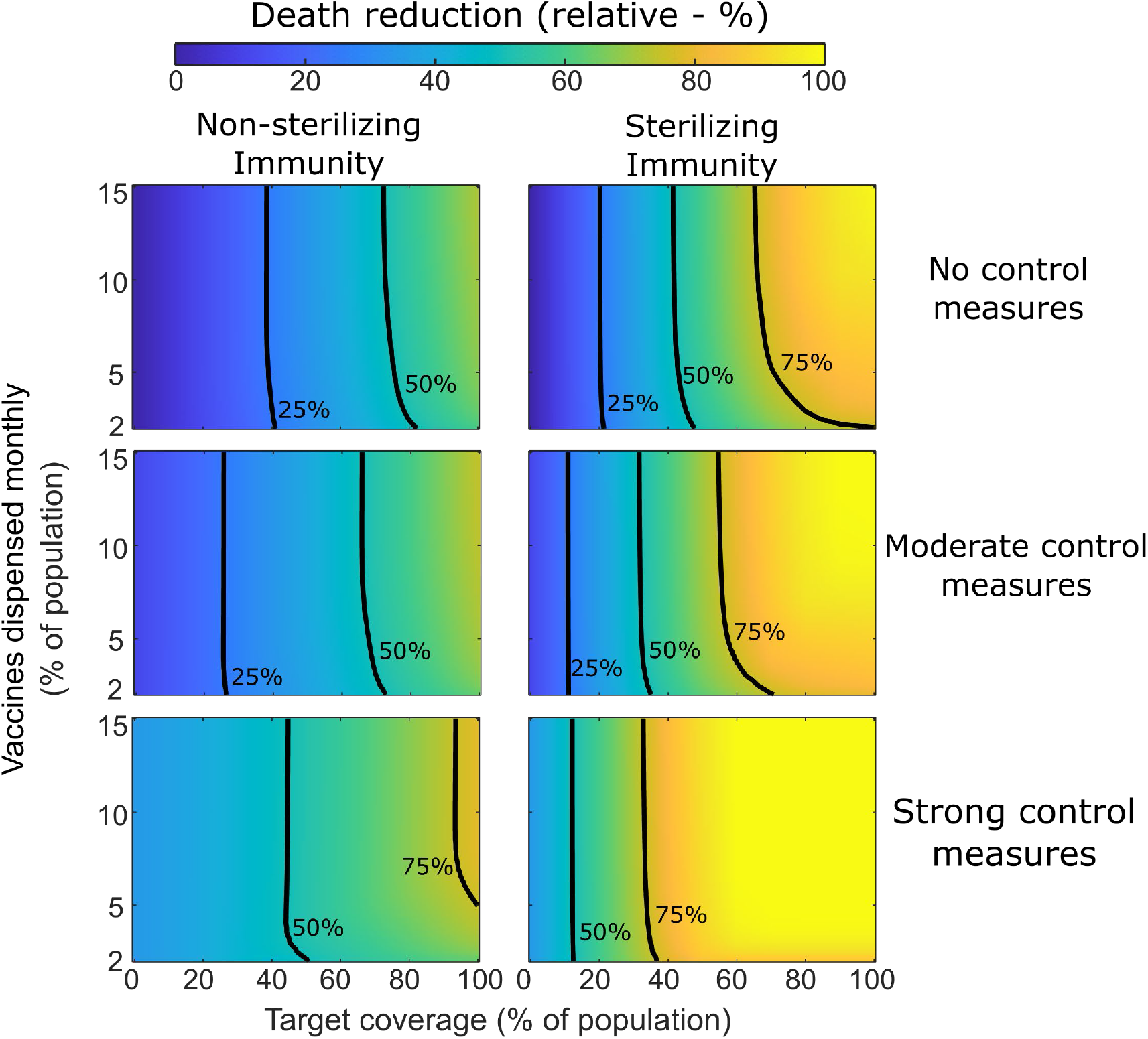
Relative reduction in deaths using vaccination strategy four. Effectiveness of strategy 4 comparative to no vaccination and no control measures is given under varying dispensation speeds, and to different maximum population coverage levels, with and without control measures. Contour lines represent 25%, 50% and 75% reductions in cumulative deaths, comparative to no vaccination and no control measures. All simulations were performed using an *R*_0_ of 2.4.

## DISCUSSION

The findings of our models support international recommendations to prioritize COVID-19 vaccine allocation for older adults (World Health Organization 2020b). Our model indicated that prioritizing vaccine allocation in older populations will contribute to the greatest relative reduction in mortality in all scenarios considered in our model. Our analyses indicate that prioritising younger populations will have a greater impact on reducing incidence of infections relative to prioritizing older age groups.

However, this reduction is marginal and prioritizing younger age groups will contribute the lowest relative reduction on COVID-19 mortality compared to other vaccine allocation strategies, including equal distribution to the general population. These findings were consistent, although to different degrees, across all model iterations, including COVID-19 vaccines that confer sterilizing and non-sterilizing immunity. A similar framework for equitable allocation of COVID-19 vaccine that prioritised older populations was adopted by the panel of experts from the Centers for Disease Control and Prevention (CDC), the National Institutes of Health (NIH) and the National Academies (Gayle et al. 2020).

The characteristics of first-generation COVID-19 vaccines remain a question of debate (Peiris and Leung 2020). However, it is unlikely that leading vaccine candidates will provide 100% protective efficacy or protection from asymptomatic carriage. For this reason, we varied vaccine efficacy and the type of immune response conferred by COVID-19 vaccines in our models. Most candidate vaccines that are currently in Phase III trials aim to assess efficacy against clinical disease (Peiris and Leung 2020). Recent reports of preliminary analyses from leading COVID-19 vaccine candidates in advanced phases of clinical development indicate vaccine efficacy against confirmed cases of >90%, including among older populations (Callaway 2020). The WHO has indicated that a successful vaccine should be 50% efficacious with the lower bound of 95% confidence intervals not crossing below 30% (Krause et al. 2020). We observed greater differences between COVID-19 vaccine allocation strategies at higher vaccine efficacy values for relative reductions in deaths. Vaccines that confer sterilizing immunity led to greater relative reductions of cases and deaths compared to vaccine that did not provide sterilizing immunity. This is attributable to the fact that sterilizing vaccines contribute to disrupting transmission of the virus. However, it should be noted that COVID-19 vaccine challenge studies in primates have demonstrated reductions in symptomatic disease and viral load, but did not produce sterilising immunity (Corbett et al. 2020; van Doremalen et al. 2020). Some first-generation COVID-19 vaccines that reduce disease severity, might not effectively reduce SARS-CoV-2 transmission in humans.

Policy makers around the world, especially those in low- and middle-income countries, have had to make difficult decisions related to the implementation and relaxation of lockdown measures. Lockdown measures help to reduce transmission of the virus but have been associated with several secondary effects, including reduced livelihoods (Walker et al. 2020), increased morbidity and mortality due to limited utilization of routine health services (Roberton et al. 2020), and several psychosocial and mental health implications (Roy et al. 2020). Effective COVID-19 vaccines are viewed as one intervention that could alleviate the need for restrictive lockdown measures. Our model allowed us to make relative comparisons of COVID-19 vaccine allocation strategies in the context of various control measure scenarios. We found that the relative reduction in cases and deaths does not meaningfully change based on the level or absence of control measures when the vaccine does not provide sterilizing immunity. However, in the model where effective vaccines do provide sterilizing immunity, the relative reduction in cases and deaths is substantially greater when strong control measures are in place and minimizes the effect of vaccine dose availability.

In accordance with COVAX Facility requirements (World Health Organization 2020a), the allocation of COVID-19 vaccines to health care workers and social workers will need to be prioritized. There remains an acute health workforce shortage in many parts of the country (Rao et al. 2016; Shrivastava and Shrivastava 2019). Immunizing this important population with priority, together with adequate supply of personal protective equipment, will help to strengthen the resiliency of the fragile health system during the epidemic. While health workers remain at higher risk of SARS-CoV-2 infection (Nguyen et al. 2020), there is insufficient evidence to determine whether they contribute to transmission greater than other subgroups within any population. Our model, did not consider a health worker compartment and therefore does not make any assumptions about the contribution of this population to overall transmission of COVID-19 in India.

With regard to the introduction of new health interventions, it is imperative to consider the experience of previous pandemics. During the 2009 H1N1 influenza pandemic, two issues that were raised were the need for accurate supply forecasts to inform vaccine ordering and subsequent distribution and the need for clear communication about priority groups for vaccination (Rambhia et al. 2010). Population structure was an important factor in determining the optimal vaccine distribution (Lee et al. 2010; Matrajt and Longini Jr 2010). Similarly, researchers have observed age-specific transmission dynamics and clinical features associated with SARS-CoV-2 (Bi et al. 2020; Laxminarayan et al. 2020). Our age-structured model is able to account for these factors.

It has also traditionally been considered important that the elderly and those particularly vulnerable to infection (e.g., those with chronic respiratory diseases) be given priority while considering influenza vaccination strategies. However, targeting the elderly in vaccination policies has not maximally reduced influenza-related mortality rates (Baguelin et al. 2013). Vaccine efficacy has also been found to decrease with age. A pooled analysis of vaccine efficacy over five influenza seasons among adults greater than 18 years who were systematically enrolled in the U.S. Flu Vaccine Efficacy Network showed that vaccine efficacy against A(H3N2) viruses was 14% (95% confidence interval [CI]: 14-36%) for adults ≥65 years versus 21% (95% CI: 9%-32%) for adults 18–49 years (Russell et al. 2018). Vaccinating high transmission groups who tend to become infected more frequently, remain infected for longer, and therefore acting as vectors to bring the infection to the elderly had been found to be more successful (Baguelin et al. 2013). However, adapting this to the COVID-19 scenario would depend on the immunogenicity and immune correlates of specific potential COVID-19 vaccines, which are currently unknown.

India has a robust national immunization program for early childhood that has been strengthened recently with demonstrable and striking gains in vaccination coverage (Gurnani et al. 2018). The recent introduction and rollout of the pneumococcal conjugate vaccine and rotavirus vaccine have shown that new vaccines can be successfully rolled out within existing public health infrastructure (Malik et al. 2019). While a clear strategy for childhood vaccination exists globally and in India, a blueprint for adult immunization is recognizably inadequate and is being increasingly acknowledged as important for sustaining and enhancing health outcomes, as well as social and economic outcomes (Privor-Dumm et al. 2020b). The framework for vaccine delivery for older adults depends on factors such as availability of evidence, existing vaccination policies, political will, robust surveillance systems, funding allocation, and importantly, communication and public messaging (Privor-Dumm et al. 2020a). India has initiated the process of targeting adults with setting up of health centers adult and immunization as an example of a life course approach to health services (Lahariya and Bhardwaj 2020).

Previous experiences with vaccine deployment in pandemic settings provide several lessons learned that may be utilized to enhance current allocation plans for a COVID-19 vaccine. During the influenza pandemic of 2009, the WHO Initiative was able to deploy almost 80 million doses of pandemic H1N1 vaccine to resource-limited settings in 77 poorest countries (World Health Organization 2012). Experiences from Latin America and the Caribbean during the influenza vaccine rollout a decade ago indicated that despite having a national preparedness plan in place and building on existing immunization infrastructure, there were inequities in vaccine distribution especially amongst the most at-risk populations (Ropero-Álvarez et al. 2016). Lessons for future vaccine deployment in emergency situations include accurate estimation of some high-risk groups and prioritizing risk-based vaccine allocation. First, the availability of robust evidence of demographics, including at-risk population groups is critical for successful vaccine utilization. The utility of simulations with varying scenarios such the current report can complement evidence and play an important role in allocation decisions. Second, coordinated planning of national vaccine deployment including establishment of a robust supply chain management system was crucial to effective utilization of scarce vaccine resources. Third, funding support from world agencies, local funders and governments can help sustain the vaccine rollout. Finally, public communication and clear messaging is essential to enhancing public confidence in vaccines.

Due to data availability constraints, and evolving scientific understanding of COVID-19, the model makes a number of key assumptions about transmission dynamics. Infection was assumed to provide temporary immunity against reinfection for one year, with other values explored in the supplemental materials. The actual average length of immunity due to COVID-19 infection is not precisely known and likely varies based on infection severity (Randolph and Barreiro 2020; van der Heide 2020; Wajnberg et al. 2020). Many model parameters, such as force of infection, latent period, time to recovery, and vaccine efficacy all likely vary with age, and potentially with time. However, given lack of clear data, these factors were assumed constant. In addition, due to data availability, deaths were estimated using case fatality ratios and not infection fatality ratios, under the assumption that discrepancies between case and infection fatality ratios are predominantly due to undetected asymptomatic infections.

Within the model, vaccines were distributed to a target coverage level, which was constant for each age group. For practical implementation, certain age groups will likely be easier to reach and less reticent to vaccination than others, meaning true coverage may vary by age (Cobos Muñoz et al. 2015). Preliminary evidence suggests that COVID-19 may be subject to seasonal forcing (Sajadi et al. 2020). This was not accounted for in the model to lack of a clear timeline for when vaccine rollout would begin. Given current understanding of COVID-19 immunity dynamic, there will likely be some prevalence of infection-driven immunity that exists before vaccine rollout begins. However, given uncertainties associated with vaccine delivery timelines, expected seroprevalence estimates, and the quality and duration of immunity from natural infection, there is no reliable data to inform this within the model. As a result, no prior immunity within the population was assumed. More broadly, this model was designed for comparison between vaccination strategies, and is not meant provide exact estimates of cumulative deaths or symptomatic infections. Rather results are meant to represent the estimated relative benefit of different scenarios.

## CONCLUSIONS

Progress towards development and approval of SARS-CoV-2 vaccines has been extraordinarily fast, however challenges of fair and optimal allocation remain. Supply limitations and logistic challenges suggest that vaccine administration across India will be slow, necessitating distribution strategies which offer the greatest protection. Within this study we have illustrated that when accounting for Indian population structure, vaccination of older age groups (>60yrs old) consistently provides the greatest reduction in cumulative deaths. Prioritized vaccination of younger age groups was often seen to reduce symptomatic infection rates, but this benefit was typically offset by the much larger infection fatality rates in older populations. Prioritized vaccination of older populations was seen to be optimal regardless of vaccine efficacy, dispensation speed, force of infection and target coverage, and independent of whether non-pharmaceutical interventions were implemented.

## Data Availability

The authors confirm that the data supporting the findings of this study are available within the article and its supplementary materials.

## Conflict of interest

The authors report no conflicts of interest.

## Sources of support

No funding or other support was received for this work.

## Author contributions

CB conceptualized the project and collated the data. BF and BW developed the model and developed the code. BF prepared the visualizations. All authors contributed to the design of the model, the interpretation of the results, and the initial draft of the manuscript.

## Ethical approvals

No human subjects were involved in this work and therefore ethical approvals were not required for the development of this manuscript.

## Notes

### Competing Interest Statement

The authors have declared no competing interest.

### Funding Statement

No external funding was received for this work.

### Author Declarations

No IRB approval was needed for this study since it involves mathematical modelling using data available in the public domain.

