## Supplemental material for "Comparing COVID-19 vaccine allocation strategies in India: a mathematical modelling study"

**SUPPLEMENTAL FILE**

### Comparing COVID-19 vaccine allocation strategies in low- and middle-income countries: a mathematical modelling study using data from India

#### 1.0 Example fit for estimate of R0

Simulations within this study were performed using three estimates of R0 (2.40, 1.81, 3.74) representing the mean, minimum and maximum estimated R0 values over the 10 most populous regions of India (data given in **Table 1**). For illustrative purposes, in **eFigure 1** we present the model fit for one of these regions – Karantaka. Note that the model compartment Q (self-isolation/quarantined/hospitalized population) was fit against the confirmed active case numbers for the given region. This is because confirmed active cases are likely to adhere to self-isolation or were diagnosed through hospitalisation.

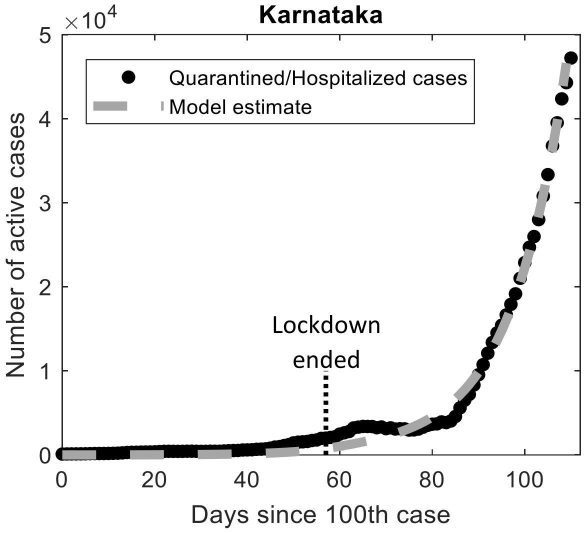

**eFigure 1 – Model fit used for estimate of R0 in Karnataka.** Comparison between the total number of active confirmed cases and simulated number of confirmed (quarantined/hospitalized) cases is given over time. Time is presented relative to the number of days since the $100^{th}$ case, and the time point at which lockdown ended (May $31^{st}$, 2020) is annotated.

#### 2.0 Simulations with varying R0 values

Within the main manuscript we presented results for simulations using an R0 value of 2.40, representing the mean estimated R0 for the 10 most populous regions of India. For completeness here we present similar results using low (1.81) and high (3.74) values of R0. These values represent the minimum and maximum R0 estimate from the same 10 regions (R0 estimates given in **Table 1** of the main manuscript). In **eFigure 2** and **eFigure 3** we present simulation results for low and high R0 values respectively, for both sterilizing and non-sterilizing vaccines. Similar to in the main manuscript, results are presented under multiple control measures, vaccine efficacies and rollout speeds, for the four main strategies.

As in the main manuscript, strategy 4 (giving vaccination priority to subjects > 60 yrs old) is seen to provide the greatest reduction in deaths, regardless of control measures, efficacy, rollout speed or whether the vaccine is sterilizing or not. There is no consistent optimal strategy for reducing symptomatic infections, with relative strategy performance varying with control measures, efficacy, vaccine type, and the R0 value.

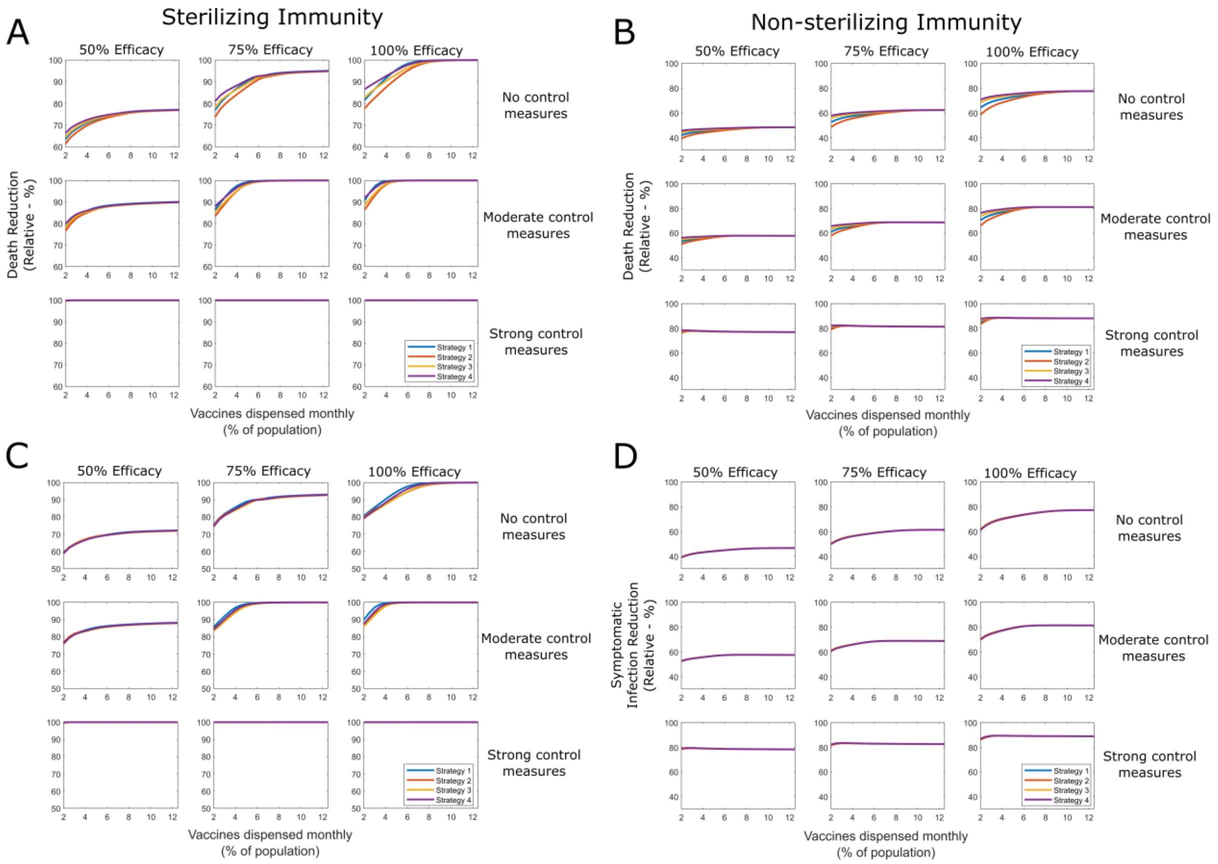

**eFigure 2 – Reductions in deaths and symptomatic infections for sterilizing and non-sterilizing vaccination strategies assuming a low R0 (1.81).** Simulations were performed using no, moderate and strong control measures, and vaccine efficacies of 50%, 75% and 100%. For both vaccine types, Strategy 4 consistently results in the greatest death reductions, while the optimal strategy for reducing symptomatic infections is Strategy 1 (sterilizing vaccine), or is dependent on rollout speed, control measures and efficacy (non-sterilizing vaccine). Simulations were performed using 75% target coverage.

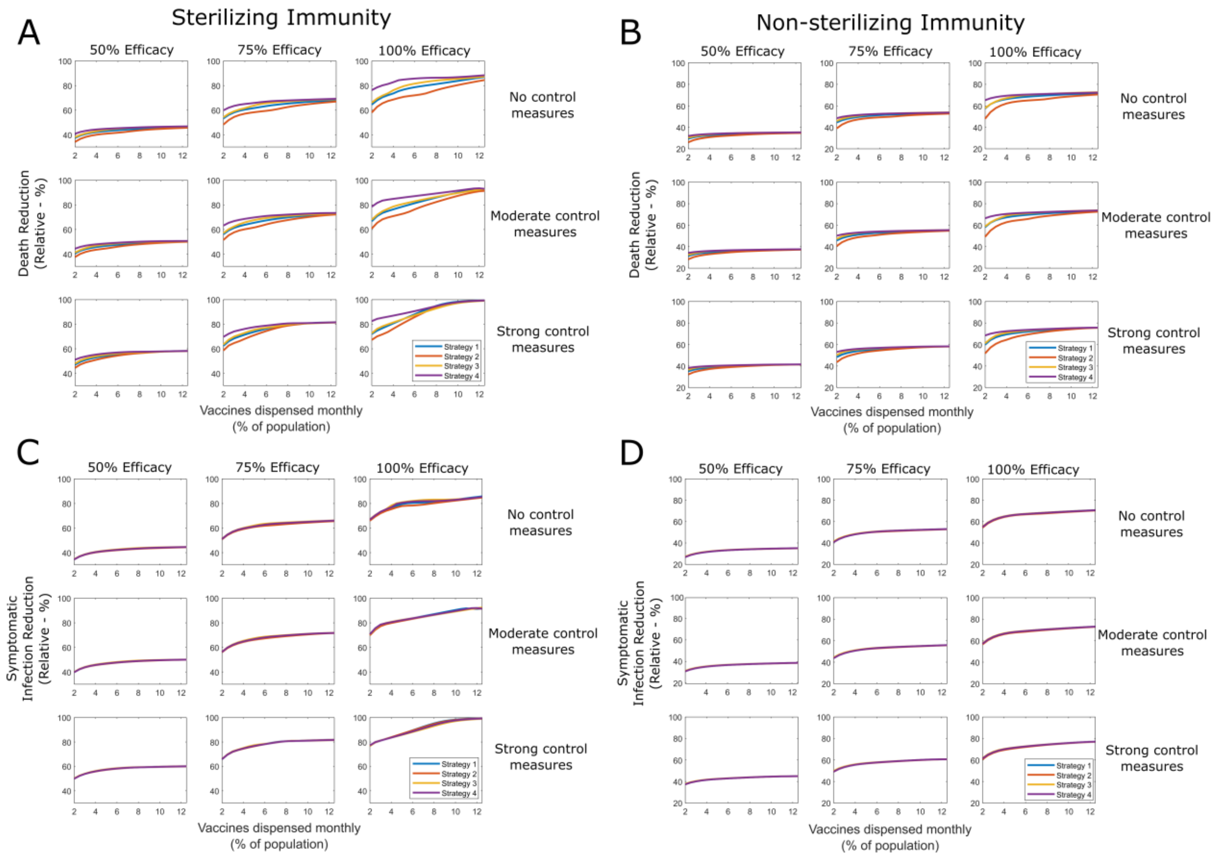

**eFigure 3 – Reductions in deaths and symptomatic infections for sterilizing and non-sterilizing vaccination strategies assuming a high R0 (5.00).** Simulations were performed using no, moderate and strong control measures, and vaccine efficacies of 50%, 75% and 100%. For both vaccine types, Strategy 4 consistently results in the greatest death reductions, while the optimal strategy for reducing symptomatic infections was dependent on rollout speed, control measures, efficacy and whether the vaccine was sterilizing or not. Simulations were performed using 75% target coverage.

#### 3.0 Simulations with varying immunity lengths

Within the main manuscript we assumed that immunity to reinfection lasted for an average of 1 year post the initial infection. Given the recency of the emergence of SARS-CoV-2, there are not yet clear estimates of the average length of time an infected subject retains immunity to reinfection. To account for this uncertainty, in **eFigure 4** and **eFigure 5** we present results assuming a 6 month and 2 year immunity period respectively.

In both cases, Strategy four consistently results in the greatest reduction in deaths. When assuming a long immunity period (2 years), under certain conditions, small increases in the vaccines dispensed per month appear to lead to increased deaths. This is because if the infection-driven immune period is long enough, then deaths attributed to an initially severe wave of infections may be offset by the increased immunity they create, generating a degree of natural (non-vaccine) herd immunity (this scenario is illustrated in **eFigure 6**). In practice, the exact immune dynamics generated by SARS-CoV-2 are not precisely known and likely vary with age, meaning natural herd immunity is not in any way reliable. Equally, in the overwhelming majority of cases, increases in vaccine distribution speed lead to significant reductions in deaths.

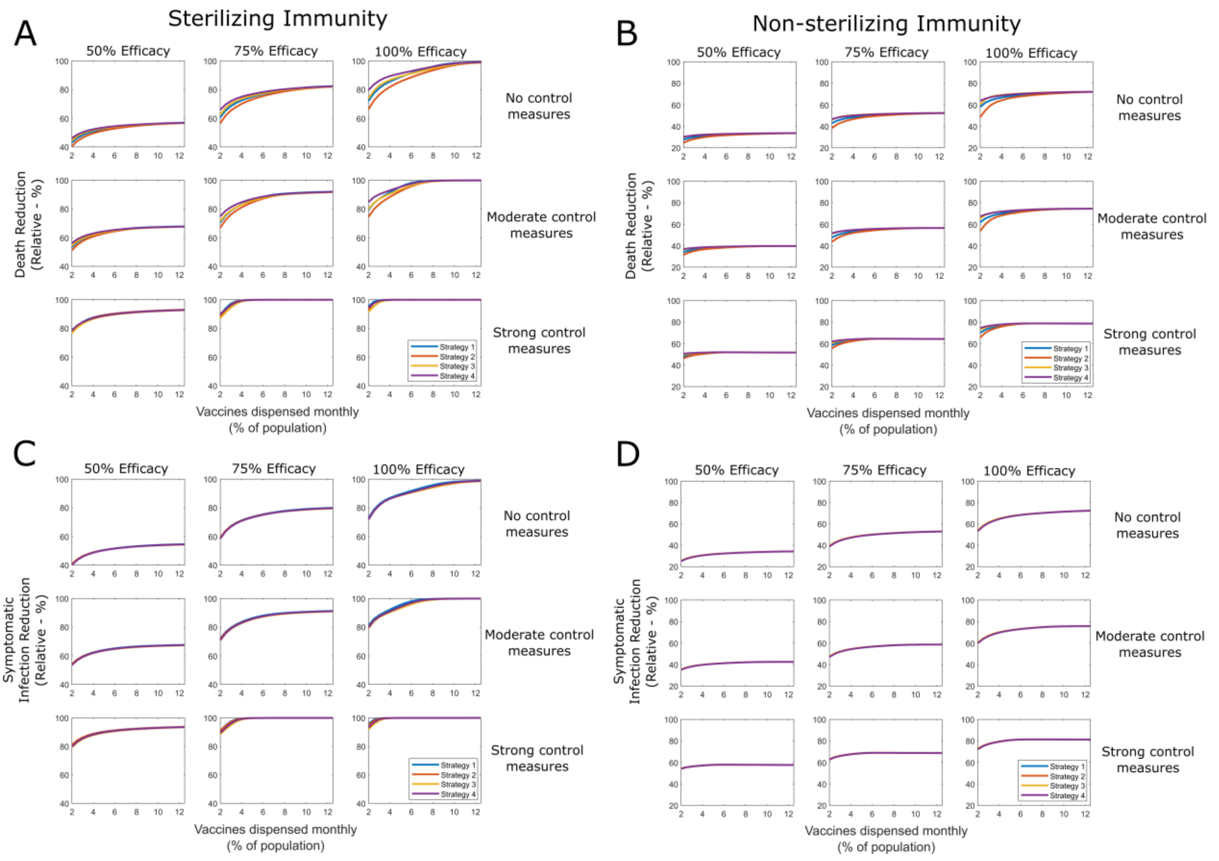

**eFigure 4 – Reductions in deaths and symptomatic infections for sterilizing and non-sterilizing vaccination strategies assuming a short immunity period.** Simulations were performed using no, moderate and strong control measures, and vaccine efficacies of 50%, 75% and 100%. Infection-driven immunity to reinfection was assumed to last 6 months. For both vaccine types, Strategy 4 consistently results in the greatest death reductions, while the optimal strategy for reducing symptomatic infections was dependent on rollout speed, control measures, efficacy and whether the vaccine was sterilizing or not. Simulations were performed using a 75% target coverage.

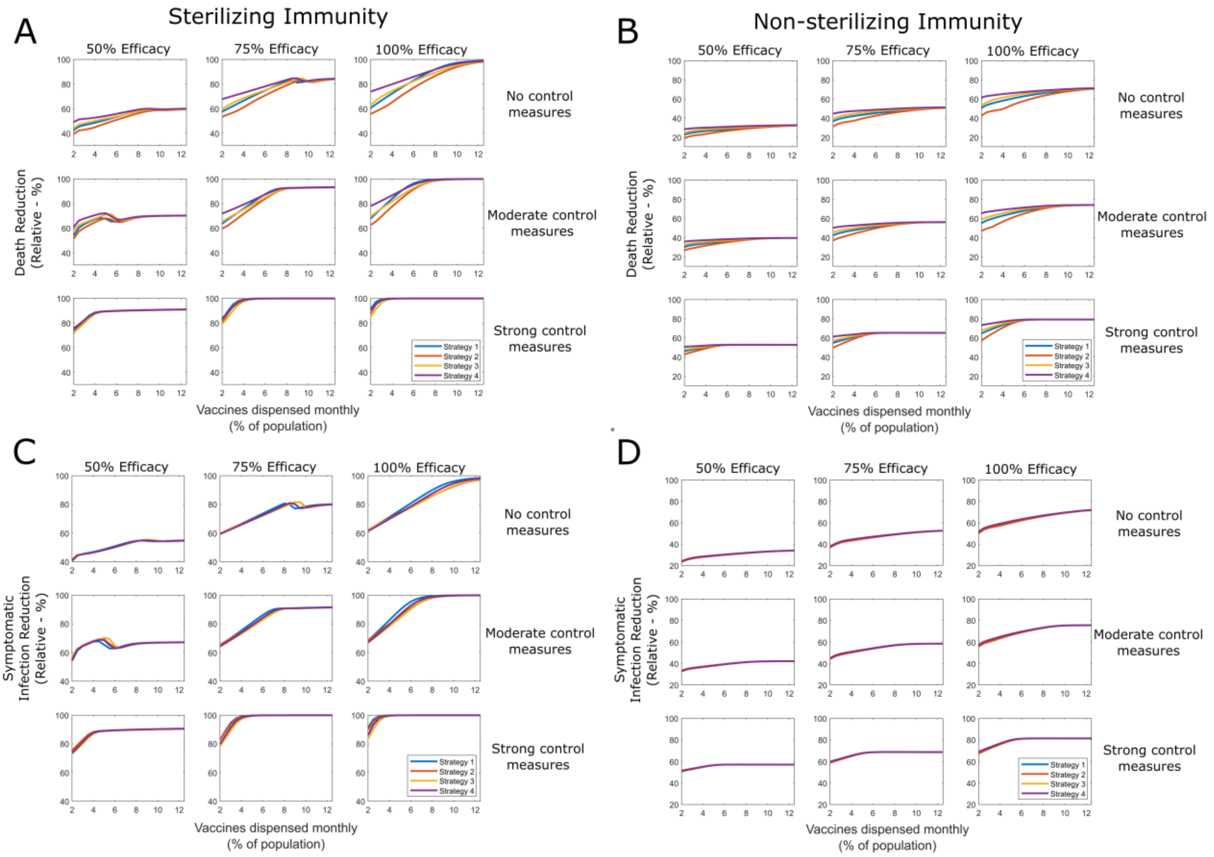

**eFigure 5 – Reductions in deaths and symptomatic infections for sterilizing and non-sterilizing vaccination strategies assuming a long immunity period.** Simulations were performed using no, moderate and strong control measures, and vaccine efficacies of 50%, 75% and 100%. Infection-driven immunity to reinfection was assumed to last 2 years. For both vaccine types, Strategy 4 consistently results in the greatest death reductions, while the optimal strategy for reducing symptomatic infections was dependent on rollout speed, control measures, efficacy and whether the vaccine was sterilizing or not. Simulations were performed using a 75% target coverage. At a few key points increases in vaccine dispensation speed result in increased deaths, due to the effects of natural immunity generated by severe infection waves.

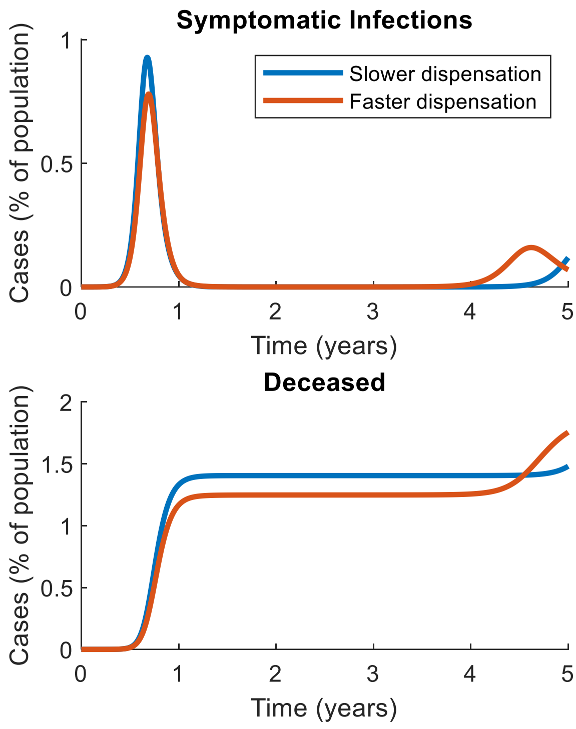

**eFigure 6 – Illustration of the effect of natural immunity against small changes in dispensation speed.** When assuming a long natural immunity period (2 years), paradoxically, small increases in vaccine dispensation speed may lead to small increases in total deaths. This is because of natural herd immunity, whereby a more severe early infection wave may create more widespread immunity, increasing the length of time until the next wave. Given uncertainties in immunity dynamics of SARS-CoV-2 this phenomenon cannot be reliably predicted and should not inform policy decision making. Simulations were performed using an R0 of 2.40, vaccine efficacy of 75%, moderate control measures and a target coverage of 75%. Slower dispensation was assuming 5% of the population was vaccinated per month, while faster dispensation assumed 6% were vaccinated per month.

<u>4.0 Simulations with varying target coverage levels</u>

Within the main manuscript results in **Figure 3** assumed a target coverage of 75%. For completeness in **eFigure 7** and **eFigure 8** we present similar results using coverages of 25% and 50%. Broadly consistent results can be seen, with Strategy 4 consistently leading to the greatest reduction in deaths. As target coverage is lowered, the overall reduction in deaths is decreased, and relative differences in effectiveness of each strategy are reduced.

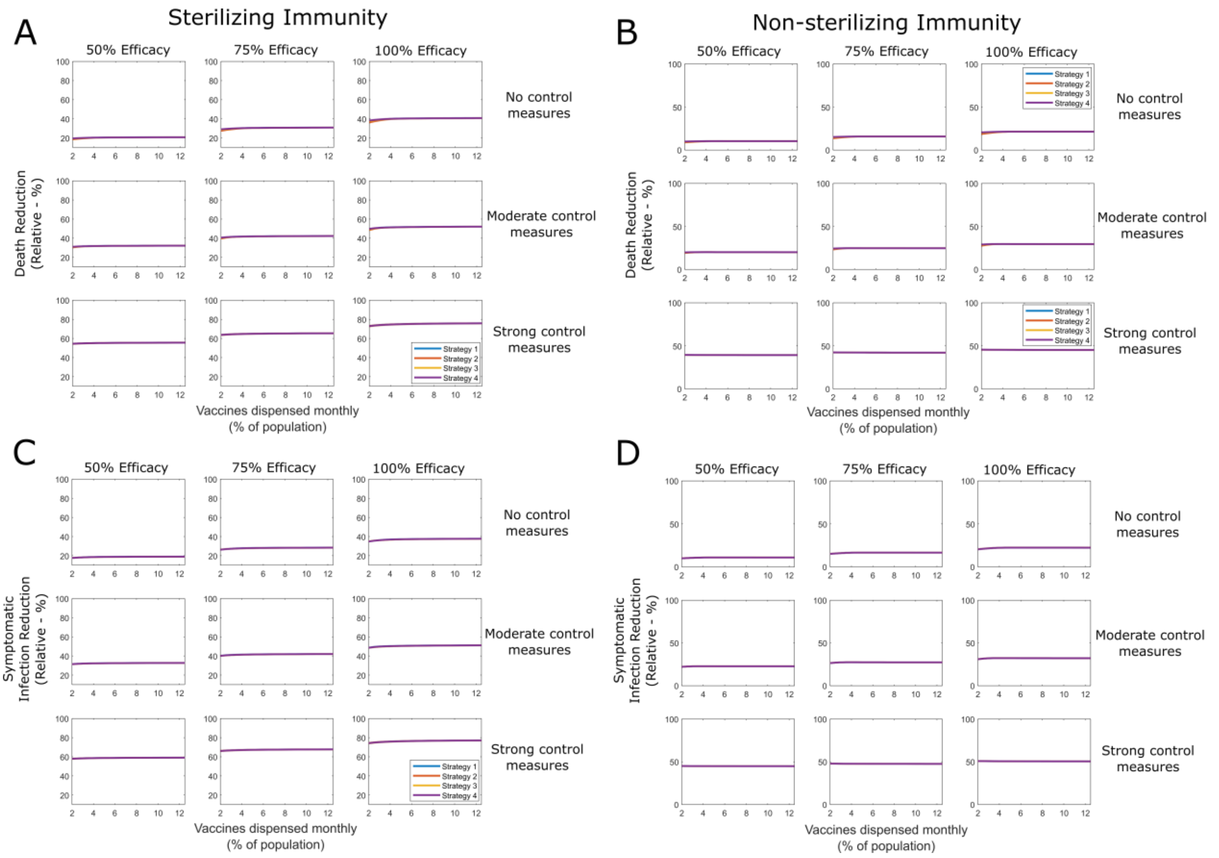

**eFigure 7 – Reductions in deaths and symptomatic infections for sterilizing and non-sterilizing vaccination strategies assuming a target coverage of 25%.** Simulations were performed using no, moderate and strong control measures, and vaccine efficacies of 50%, 75% and 100%. Infection-driven immunity to reinfection was assumed to last 2 years. For both vaccine types, Strategy 4 consistently results in the greatest death reductions, while the optimal strategy for reducing symptomatic infections was dependent on rollout speed, control measures, efficacy and whether the vaccine was sterilizing or not.

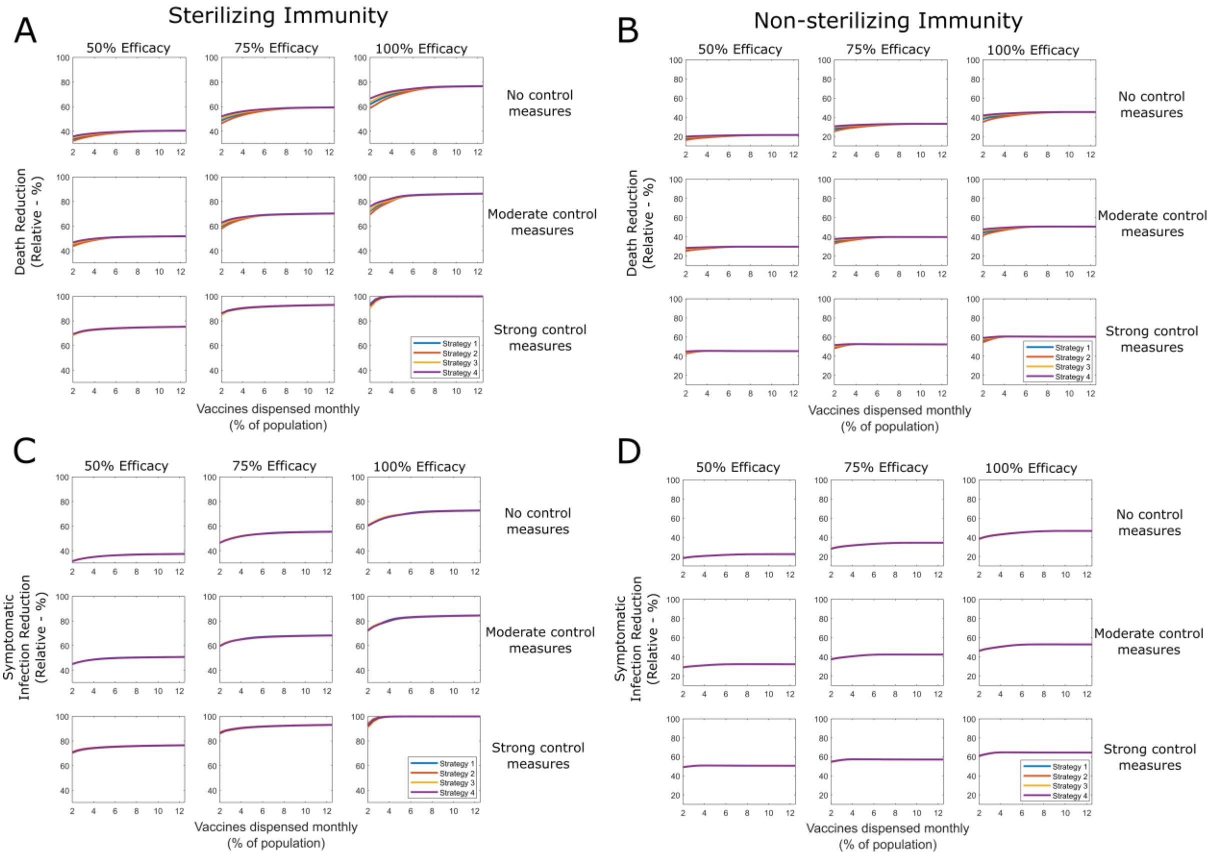

**eFigure 8 – Reductions in deaths and symptomatic infections for sterilizing and non-sterilizing vaccination strategies assuming a target coverage of 50%.** Simulations were performed using no, moderate and strong control measures, and vaccine efficacies of 50%, 75% and 100%. Infection-driven immunity to reinfection was assumed to last 2 years. For both vaccine types, Strategy 4 consistently results in the greatest death reductions, while the optimal strategy for reducing symptomatic infections was dependent on rollout speed, control measures, efficacy and whether the vaccine was sterilizing or not.

#### 5.0 Simulations with imperfect self-isolation

The model in the main manuscript assumes that eventually symptomatically infectious individuals will self-isolate or be hospitalised after which they will no longer transmit infection to others. However, while most individuals will likely adhere to self-isolation or quarantine practices when actively symptomatic, in practice self-isolation will never be perfectly adhered to. To illustrate this effect, we update the model to incorporate imperfect self-isolation, where individuals in this compartment can still infect susceptible individuals, albeit at a lower rate. Ignoring vaccination and waning immunity contributions, infection-driven changes to the susceptible population is expressed as

$$\frac{dS_i}{dt} = -\frac{\beta_1}{N}\sum_{j=1}^{K} C_{ij}\, S_i\, \left(A_j + A_j^v + I_j\right).$$

To account for an imperfect self-isolation, we update this term as

$$\frac{dS_i}{dt} - \frac{\beta_1}{N}\sum_{j=1}^{K} C_{ij}\, S_i\, \left(A_j + A_j^v + I_j + fQ_j\right),$$

where *f* is the adherence to self-isolation (with *f* = 1 representing perfect adherence, and *f* = 0 representing no adherence). Equivalent updates are made to the appropriate terms in the $V_i, E_i$, and $E_i^v$ differential equations in the main manuscript.

Within **eFigure 9** we present an equivalent of **Figure 3** from the main manuscript, using an imperfect self-isolation, with adherence of 0.9, an $R_0$ of 2.40 and a population coverage of 75%. Qualitatively similar results to **Figure 3** can be seen, with strategy 4 consistently resulting in the greatest relative reduction in deaths, with less discrepancy across the four strategies in reducing symptomatic infections.

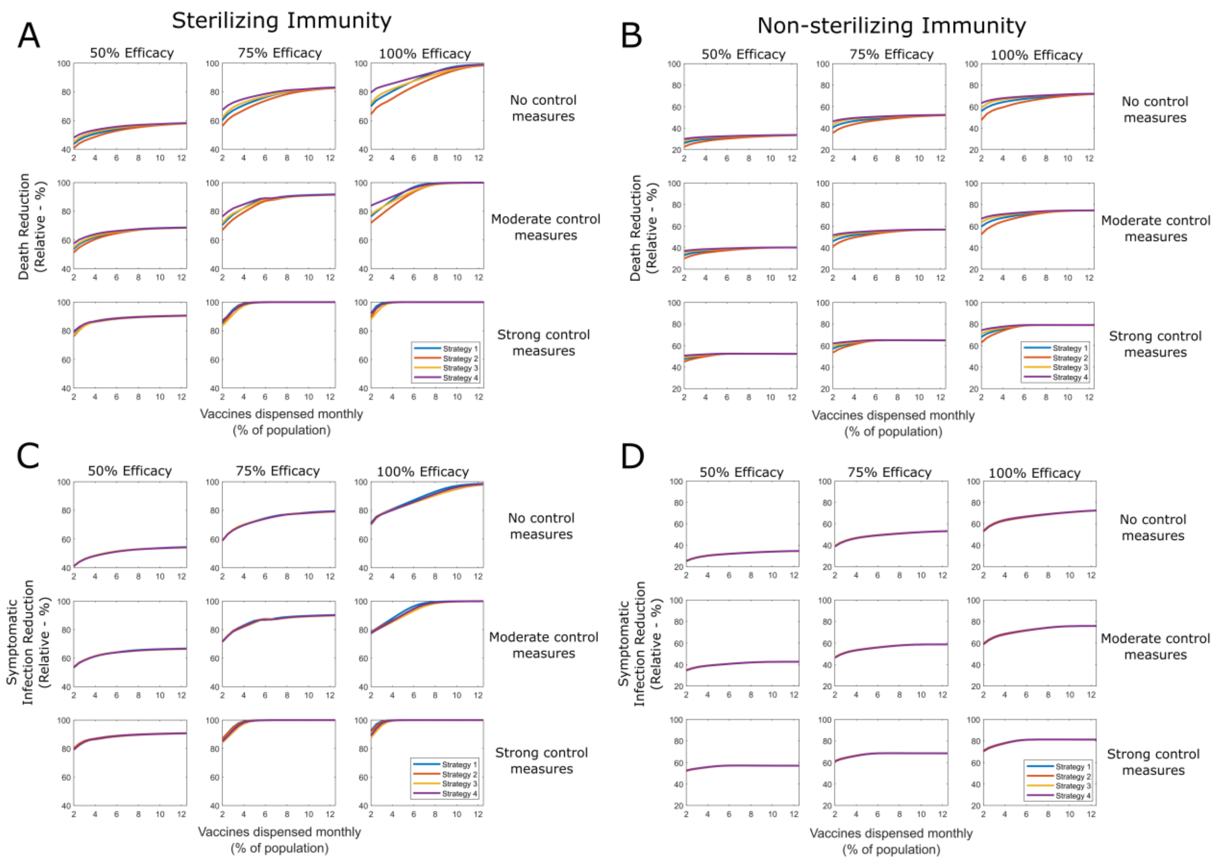

**eFigure 9 – Reductions in deaths and symptomatic infections for sterilizing and non-sterilizing vaccination strategies assuming imperfect self-isolation.** Simulations were performed using no, moderate and strong control measures, and vaccine efficacies of 50%, 75% and 100%. Adherence to self-isolation was assumed to be 90% ($f$ = 0.9). For both vaccine types, Strategy 4 consistently results in the greatest death reductions, while the optimal strategy for reducing symptomatic infections was dependent on rollout speed, control measures, efficacy and whether the vaccine was sterilizing or not.

6.0 Illustration of coverage and dispensation speed dynamics for vaccination strategies 1-3

Within the main manuscript in **Figure 4** we presented heatmaps illustrating the relative reduction in deaths using strategy 4 under variations in dispensation speed and target coverage. For completeness in **eFigures 10-12** we present similar results for strategies 1, 2 and 3 respectively. Quantitatively similar results can be seen to **Figure 4**, though with overall slightly lower reductions in deaths than equivalent results for strategy 4.

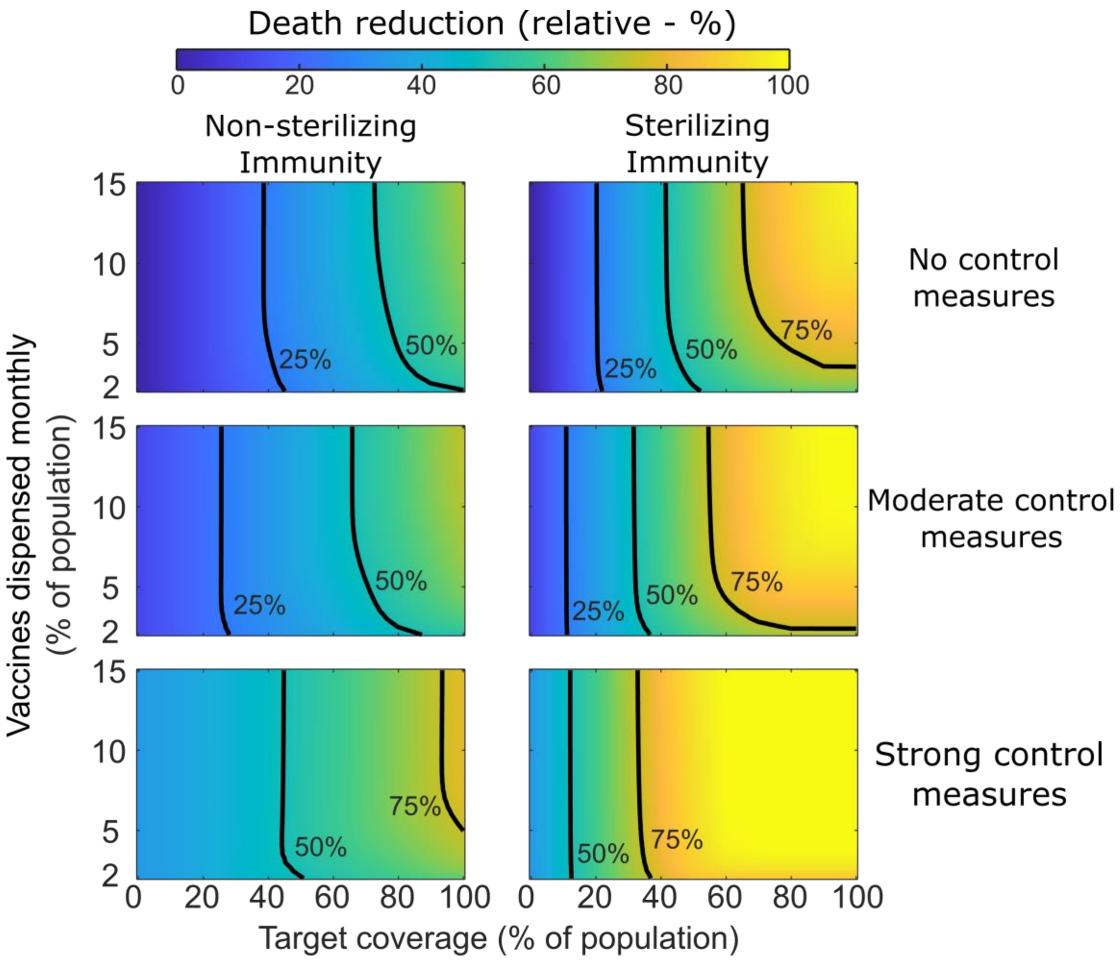

**eFigure 10 – Relative reduction in deaths using vaccination strategy four.** Effectiveness of strategy 1 comparative to no vaccination and no control measures is given under varying dispensation speeds, and to different maximum population coverage levels, with and without control measures. Contour lines represent 25%, 50% and 75% reductions in cumulative deaths, comparative to no vaccination and no control measures. All simulations were performed using an R0 of 2.40.

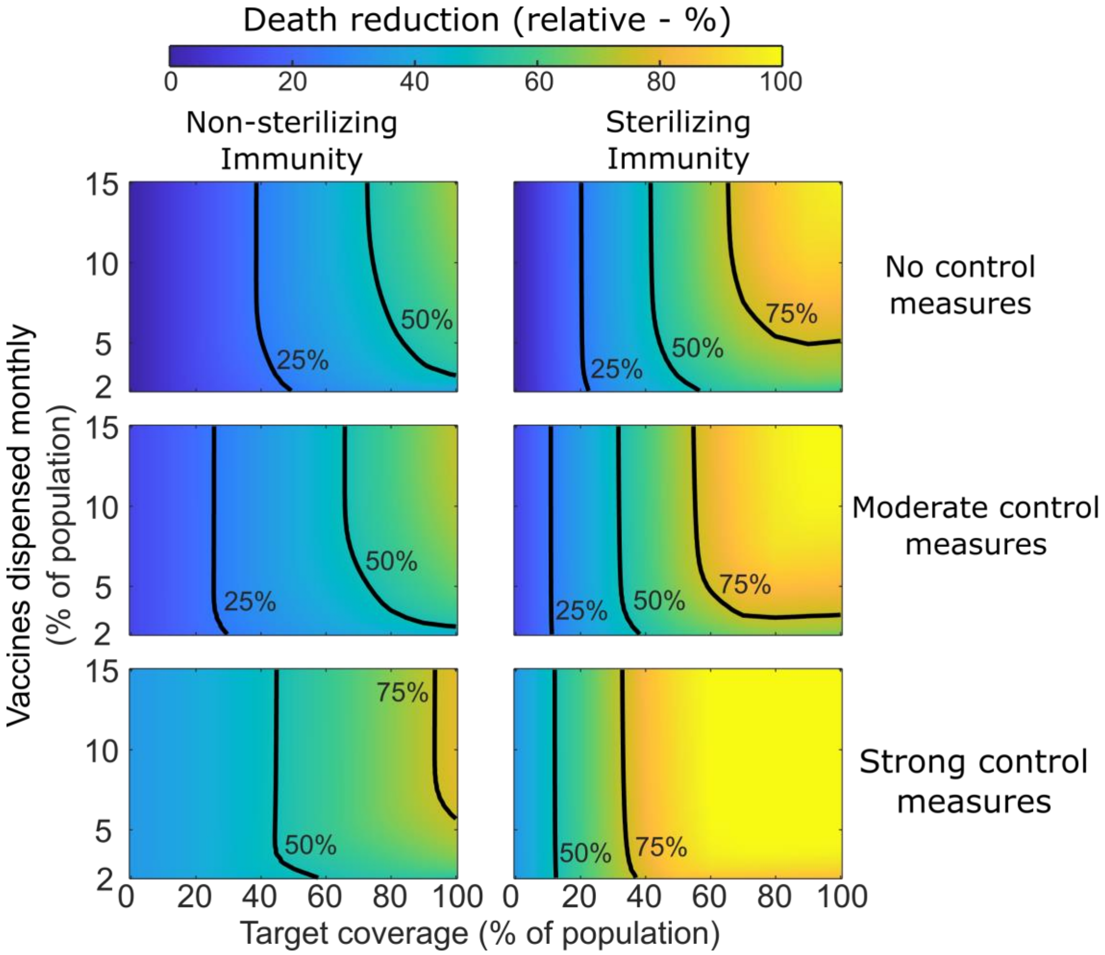

**eFigure 11 – Relative reduction in deaths using vaccination strategy four.** Effectiveness of strategy 2 comparative to no vaccination and no control measures is given under varying dispensation speeds, and to different maximum population coverage levels, with and without control measures. Contour lines represent 25%, 50% and 75% reductions in cumulative deaths, comparative to no vaccination and no control measures. All simulations were performed using an R0 of 2.40.

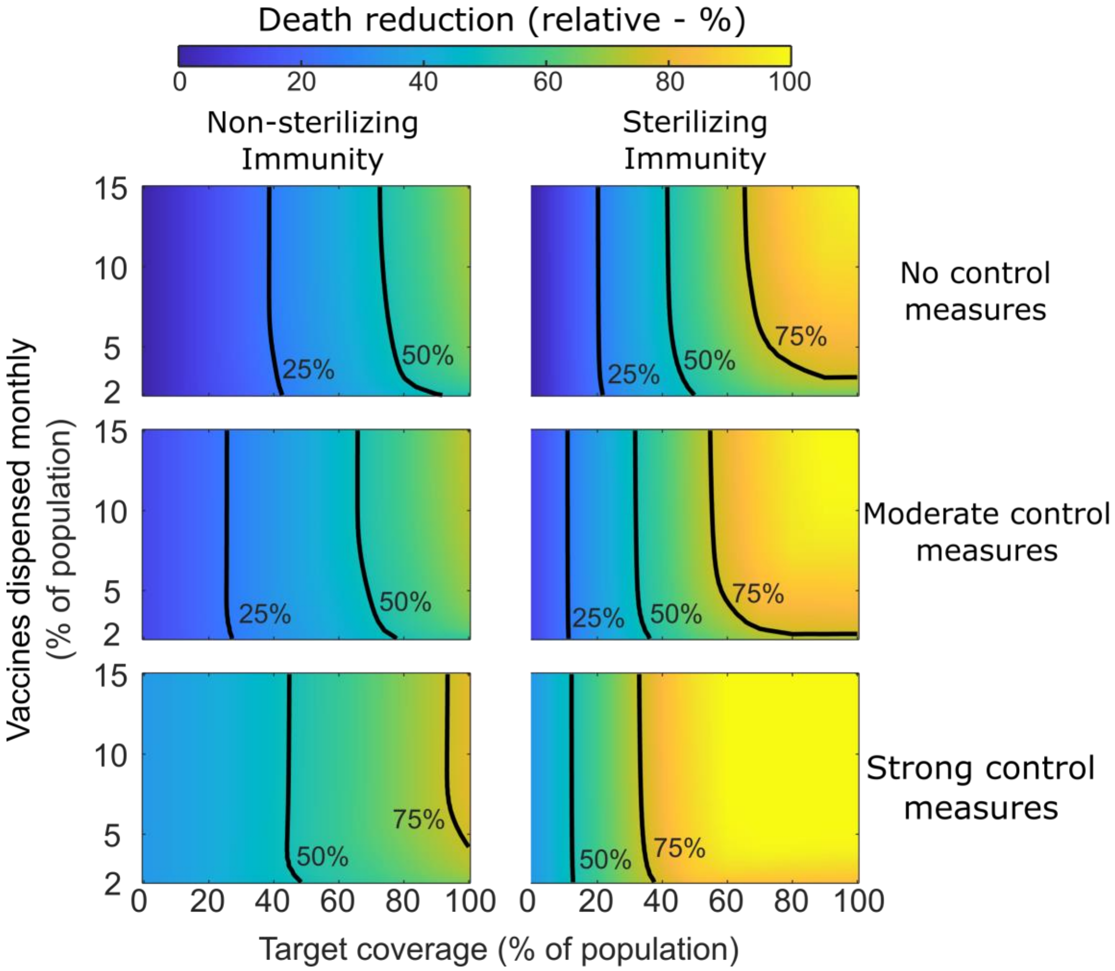

**eFigure 12 – Relative reduction in deaths using vaccination strategy four.** Effectiveness of strategy 3 comparative to no vaccination and no control measures is given under varying dispensation speeds, and to different maximum population coverage levels, with and without control measures. Contour lines represent 25%, 50% and 75% reductions in cumulative deaths, comparative to no vaccination and no control measures. All simulations were performed using an R0 of 2.40.
